# Epidemiological characteristics of an outbreak of Coronavirus Disease 2019 in the Philippines

**DOI:** 10.1101/2020.04.12.20053926

**Authors:** Mark M. Alipio, Joseph Dave M. Pregoner

**Author notes:** Contributed equally.

## Abstract

The outbreak of Coronavirus disease 2019 (Covid-2019) is a source of great concern in the Philippines. In this paper, we described the epidemiological characteristics of the laboratory-confirmed patients with Covid-2019 in the Philippines as of April 3, 2020 and provided recommendations on how to limit the spread of the disease. Data from the DOH NCOV tracker and University of the Philippines’ Covid-2019 tracker were extracted, from its initiation (January 30, 2020) until the most recent situation report (April 3, 2020). The total number of cases and deaths were stratified by sex, age, and region of the Philippines. Descriptive statistics were used to analyze the demographic profile of the confirmed cases. Case fatality rate, in percent, was calculated by dividing the total number of deaths to the total number of confirmed cases. Results revealed that a total of 3,018 cases of Covid-2019 spread were confirmed across 17 regions in the Philippines. These cases occurred over the course of 73 days through person-to-person transmission, highlighting an extremely high infectivity rate. The 144 deaths accounted for, equate to 4.51 case fatality rate, seemingly lower compared to its predecessors, severe acute respiratory syndrome (SARS) and Middle East respiratory syndrome (MERS), but higher compared to that of United States of America, Germany, mainland China, and neighboring Southeast Asian countries such as Malaysia, Singapore, Brunei, and Thailand. Of the 3,018 confirmed cases, majority were male, elderly, and diagnosed in Metro Manila region. Case fatality rates were higher in male and highest among elderly and Filipinos in the Ilocos region. With the surge on the number of cases, precautionary measures should remain a responsibility, and protocols for prevention need to be set. Adherence to infection control guidelines such as but not limited to frequently handwashing for at least 20 seconds, observing coughing etiquette, wearing of masks, and social distancing should be maintained in order to contain the disease.

## 1. Introduction

The Coronavirus Disease 2019 (Covid-2019) is a source of great concern in the Philippines. It was first confirmed to have spread in the country on January 30, 2020 in Manila, Philippines. Involved first and second cases were married couples who had travel history in China, one of which was the first confirmed fatality of the disease in the country. The first case of local transmission was reported in an area in Northern Philippines from an elderly Filipino who was frequently observed in a Muslim prayer hall. He was the first confirmed death due to the local transmission of the disease in the country.^1^

Until April 3, 2020, Covid-2019 has affected 3,018 individuals and caused 144 deaths in the Philippines, and quickly spread to over 17 regions nationwide. A total of 5,530 persons had been tested, while persons under investigation and under monitoring rose up to 1,323 and 6,321, respectively. Persons under investigation (PUIs) is a category for persons who have fever and/or cough with either a travel history in the past 14 days to areas with issued travel restrictions because of Covid-2019 and/or a history of disease exposure while persons under monitoring (PUMs) refers to individuals who have a travel history in the past 14 days to areas with issued travel restrictions because of Covid-2019 and/or a history of disease exposure. During a three-month duration of pandemic, the highest number of daily cases (538) was recorded on March 31, 2020. ^1^

Several health measures have been instigated to combat the disease outbreak. Few laboratories have opened to test for suspected cases. Among which are Research Institute for Tropical Medicine (RITM) which is located in Muntinlupa, Metro Manila and sub-national laboratories in Davao, Cebu and Northern Luzon, among others. Travel bans have been imposed to hotspot areas of the disease such as mainland China and South Korea. On March 9, Philippine President Rodrigo R. Duterte declared a public health emergency in the country after signing the Proclamation No. 922. On March 12, the Code Red Sub-Level 1 alert was raised to Code Red Sub-Level 2 alert to prevent further local transmission of the disease. As part of the concerted disease-mitigating efforts, the entire Luzon and some areas in the country have been placed in an enhanced community quarantine on March 16, after releasing a memorandum to suspend classes on March 12. In an attempt to authorize the President of the country ‘to exercise powers necessary to carry out urgent measures to meet the current national emergency related to Covid-19’, the President signed the ‘Bayanihan to Heal as One Act’ on March 25. Moreover, the Department of Health (DOH) of the country is accelerating its Covid-19 testing capacity, anticipating 120,500 kits from neighboring countries.^2^

Infection control measures and supportive treatment were the current recommendations of the World Health Organization (WHO) and the Centers for Disease Control and Prevention (CDC).^3,4^ Up to the present moment, there are no known vaccines for Covid-2019. This prompted several scientists in the world to evaluate the effectiveness of previously used treatments for SARS such as lopinavir.

Characterization of the epidemiological topographies of Covid-2019 in the country is crucial for the implementation and development of effective control strategies. In this study, we described the epidemiological characteristics of the laboratory-confirmed patients with Covid-2019 in the Philippines as of April 3, 2020. The results of the descriptive, exploratory analysis of all cases were reported. We also gave recommendations on how to limit the spread of the disease.

## 2. Methods

### 2.1 Case Data Extraction

Data from the DOH NCOV tracker (https://ncovtracker.doh.gov.ph/)^1^ and University of the Philippines’ Covid-2019 tracker (https://endcov.ph/dashboard/#)^5^ were extracted, from its initiation (January 30, 2020) until the most recent situation report (April 3, 2020). The total number of cases and deaths were stratified by sex, age, and region of the Philippines.

### 2.2 Data Analysis

Descriptive statistics were used to analyze the demographic profile of the confirmed cases. Case fatality rate, in percent, was calculated by dividing the total number of deaths to the total number of confirmed cases.

### 2.3 Ethics Approval

The ethics approval was considered exempt because all data used were sourced out from an open access Covid-2019 tracker from the Department of Health, Philippines and University of the Philippines. Nevertheless, data taken were anonymized by the governing institutions to ensure full confidentiality of information pertaining to the concerned patients.

## 3. Results

### 3.1 Descriptive Statistics

As shown in Table 1, a total of 3,018 individuals were diagnosed with Covid-2019. Of the 3,018 cases, majority were male (56.49%), aged 50-59 years (20.78%), and diagnosed in Metro Manila region (39.30%). A total of 136 deaths have occurred among 3,018 confirmed cases for an overall case fatality rate of 4.51%. Case fatality rate for males was 4.22% and for females was 2.00%. The elderly age groups (≥60 years) obtained the highest case fatality rates with the 80-89 age group obtaining the highest case fatality rate of all age groups at 15.89%. By region, Ilocos area had the highest case fatality rate at 11.11%, followed by Bangsamoro at 10.00% and Central Luzon at 6.76%.

**Table 1.** Patients, deaths, and case fatality rates, for n=3,018 confirmed Covid-2019 cases in the Philippines as of April 3, 2020.

| Baseline characteristics | Confirmed cases, N (%) | Deaths, N (%) | Case fatality rate, % |
| --- | --- | --- | --- |
| Overall | 3018 (100.00) | 136 (100.00) | 4.51 |
| Sex |  |  |  |
| Male | 1705 (56.49) | 72 (52.94) | 4.22 |
| Female | 1203 (39.86) | 24 (17.65) | 2.00 |
| Age, years |  |  |  |
| Above 90 | 8 (0.27) | - | - |
| 80–89 | 107 (3.55) | 17 (12.50) | 15.89 |
| 70–79 | 381 (12.62) | 30 (22.06) | 7.87 |
| 60–69 | 588 (19.48) | 27 (19.85) | 4.59 |
| 50–59 | 627 (20.78) | 15 (11.03) | 2.39 |
| 40–49 | 435 (14.41) | 6 (4.41) | 1.38 |
| 30–39 | 442 (14.65) | 1 (0.74) | 0.23 |
| 20–29 | 283 (9.38) | - | - |
| 10–19 | 25 (0.83) | - | - |
| Region |  |  |  |
| Metro Manila | 1186 (39.30) | 52 (38.24) | 4.38 |
| Cordillera | 13 (0.43) | - | - |
| Ilocos Region | 9 (0.30) | 1 (0.74) | 11.11 |
| Cagayan Valley | 21 (0.70) | - | - |
| Central Luzon | 74 (2.45) | 5 (3.68) | 6.76 |
| Calabarzon | 154 (5.10) | 10 (7.35) | 6.49 |
| Mimaropa | 3 (0.10) | - | - |
| Bicol Region | 3 (0.10) | - | - |
| Western Visayas | 17 (0.56) | 1 (0.74) | 5.88 |
| Central Visayas | 31 (1.03) | 2 (1.47) | 6.45 |
| Eastern Visayas | 1 (0.03) | - | - |
| Zamboanga Peninsula | 2 (0.07) | - | - |
| Northern Mindanao | 3 (0.10) | - | - |
| Davao Region | 60 (1.99) | 2 (1.47) | 3.33 |
| Soccsksargen | 1 (0.03) | - | - |
| Bangsamoro | 10 (0.33) | 1 (0.74) | 10.00 |
| For validation | 1504 (49.83) | 38 (27.94) | 2.53 |

## 4. Discussion

In this study, we described the epidemiological characteristics of the laboratory-confirmed patients with Covid-2019 in the Philippines as of April 3, 2020. Results revealed that there has been a total of 3,018 confirmed cases of Covid-2019 spread across 17 regions in the Philippines. These cases occurred over the course of 73 days through person-to-person transmission, highlighting an extremely high infectivity rate.

The 144 deaths accounted for as of April 3, 2020, equate to 4.51 case fatality rate. This is relatively lower compared to the case fatality rate of severe acute respiratory syndrome (SARS) outbreak^6^ in 2002 at 11% and of Middle East respiratory syndrome (MERS) outbreak ^7^ in 2012 at 36%. However as of April 3, the Philippine case fatality rate is slightly higher compared to United States of America (2.71%), Germany (1.50%), and even mainland China (4.08%), the countries with the greatest number of cases of Covid-2019. Nevertheless, the country’s case fatality rate is obviously lower compared to Spain (9.47%) and Italy (12.33%), among the three countries with the greatest number of cases of Covid-2019. Based on the situational reports of Covid-2019 outbreak in neighboring southeast Asian countries, Cambodia and Vietnam were the only southeast Asian countries with no reported deaths due to the disease. Meanwhile, the case fatality rates of Malaysia (1.64%), Singapore (0.50%), Brunei (0.74%), and Thailand (1.06%) were lower than that of the Philippines; however, Indonesia reported a higher fatality rate at 9.13%. Despite the lower-case fatality rate, Malaysia has slightly higher number of cases at 3,483 than the Philippines.

The study also reported that most of the confirmed cases and the highest case fatality rates belong to the elderly age groups. This is consistent with the previous studies of the Novel Coronavirus Pneumonia Emergency Response Epidemiology Team in China (2020)^8,9^. Also consistent was the finding pertaining to male individuals who had seemingly higher cases and fatality rate compared to the female group. The elderly age groups were predicted to contract the disease as these groups are one of the most vulnerable in terms of acquiring the disease and immunocompromised given their immune system capacity. Hence, special executive orders were released in the local government units in the Philippines prohibiting the elderly population from going outside their houses.

Regional analysis of the Covid-2019 outbreak reported that Metro Manila had the highest reported number of cases, followed by Calabarzon. This distribution could be attributed to the geographical nature and the population of Metro Manila. Presently, Metro Manila is the seat of the Philippine government, comprising of 16 cities. It is the second most populous and the most densely populated region of the Philippines, next to Calabarzon. Meanwhile, Ilocos area had the highest case fatality rate at 11.11%, followed by Bangsamoro at 10.00% and Central Luzon at 6.76%. Ilocos Region, occupying the north-western area of the Philippines, issued several directives to combat the daunting case fatality rate. Among which were the provisions for the preparation of health information forms, identification of quarantine areas, delineated rules for the business establishments, prohibition of the spreading of fake news, regulation of ports and borders, personal and public preventive measures, and prohibition of hoarding, reselling, and price spikes.^10^

Despite the concerted efforts in battling the Covid-2019 pandemic, precautionary measures remain a responsibility, and protocols for prevention need to be set, regardless of the relatively favourable prognosis of this disease, when compared with SARS and MERS. Parallels can be drawn to the Philippines, which only further proves that this pandemic will not end except with a collective global and national effort.

Adherence to infection control guidelines such as but not limited to frequently handwashing for at least 20 seconds, observing coughing etiquette, wearing of masks, and social distancing should be maintained in order to contain the disease, thereby decreasing the number of cases while increasing the rate of recovery which ultimately will flatten the epidemiological case curve. Filipinos who have been exposed to the virus from a known patient and showing symptoms should undergo test from the respective medical facilities, be isolated and treated. After sometime when two tests have been negative, they shall be advised to remain in an isolated area for 14 days. Similarly, those who have been exposed but showing no symptoms are hereby advised to self-quarantine for 14 days. Individuals who have fever and/or cough but have no history of exposure may also seek medical attention and implement the basic infection control guidelines. Finally, sanitization, quarantine, and isolation should be practiced fervently as it has been hypothesized in previous models that these measures are essential in combating the pandemic.^11,12^

## Data Availability

The data that support the findings of this study are derived from the following public domain resources:
https://ncovtracker.doh.gov.ph/
https://endcov.ph/dashboard/

https://endcov.ph/dashboard/

## Author Contributions

All authors equally contributed in the conceptualization of the paper, data gathering, and analyses. All authors approved the final version of the manuscript.

